# High-sensitive detection and quantitation of thyroid-stimulating hormone (TSH) from capillary/fingerstick and venepuncture whole-blood using fluorescence-based rapid lateral flow Immunoassay (LFIA)

**DOI:** 10.1101/2023.01.22.23284874

**Authors:** Samar Shurbaji, Faleh Al Tamimi, Mahmoud M. Al Ghwairi, Dayana El Chaar, Salma Younes, Amin F. Majdalawieh, GianFranco Pintus, Nader Dweik, Gheyath K. Nasrallah

**Affiliations:** Collage of Dental Medicine, Qatar University, 2713 Doha, Qatar; Department of Biomedical Science, College of Health Sciences, Member of QU Health, Qatar University, 2713 Doha, Qatar; Sciences of medical laboratory, Laboratory Analysis technologists, Al-Ahliyya Amman University, Amman 2213, Jordan; Biomedical Research Center, Qatar University, 2713 Doha, Qatar; Department of Biology, Chemistry and Environmental Sciences, College of Arts and Sciences, American University of Sharjah, 26666 Sharjah, United Arab Emirates; Department of Biomedical Sciences, University of Sassari, Sassari, Italy; Department of Research, Women’s Wellness and Research Center, Hamad Medical Corporation, Doha 3050, Qatar; Clinical and Metabolic Genetics, Department of Pediatrics, Hamad General Hospital, Hamad Medical Corporation, Doha 3050, Qatar; College of Health and Life Science (CHLS), Hamad Bin Khalifa University (HBKU), Doha 34110, Qatar

**Keywords:** TSH, Evaluation, LIFA rapid test, Finecare, Roche

## Abstract

**Background:** In the last decade, point of care testing (POCT) such as lateral flow immunoassays (LFIA) were developed for rapid TSH measurement. Most of these TSH-LFIAs are designed for qualitative measurements (i.e., if TSH values >5, or >15 IU/L) and as screening tests for primary hypothyroidism in children and adults. Serum or plasma, but not venepuncture whole-blood, or fingerstick/capillary, are usually used for accurate quantitation of TSH. Studies on performance evaluation of TSH-LFIAs POCT using venepuncture or fingerstick whole-blood are limited.

**Aim:** We aim to evaluate the performance of a new fluorescence-based LFIA and its Finecare™ fluorescent reader for quantitative measurement of TSH from a fingerstick, venepuncture whole-blood, and serum.

**Methods:** 102 fingerstick, venepuncture whole-blood, and serum samples (with normal and abnormal TSH values) were analyzed by Finecare™ Rapid Quantitative LFIA test and Roche CobasPro-c503 as a reference test.

**Results:** Using serum, when compared to CobasPro-c503 reference method, Finecare™ showed high sensitivity (90.5%) and specificity (96.3%) for diagnosis of thyroid abnormalities (<0.35 or > 4.5 mIU/L). The actual test values (mIU/L) of Finecare™ showed excellent agreement (Cohen’s Kappa=0.85) and strong correlation (r=0.93, p<0.0001) with CobasPro-c503. Using venepuncture whole-blood samples, Finecare™ showed similar results to serum with high sensitivity (95.2%), specificity (97.5%), excellent agreement (Cohen’s Kappa=0.91), and very strong correlation (r=0.95, p<0.0001) with CobasPro-c503. These results suggest that Finecare™ can be used for quantitative measurement of TSH using serum or venepuncture whole-blood. These key performance indicators were slightly decreased when fingerstick whole-blood samples were used: Sensitivity (85.7%), specificity (90.0%), good agreement (Cohen’s Kappa=0.7) and very strong correlation (r=0.9, p<0.0001) with CobasPro-c503.

**Conclusion:** Our results indicate that the Finecare™ is appropriate for TSH screening and quantitative measurement of TSH using serum samples and fingersticks, particularly in none or small laboratory settings.

**Highlights:**

- Finecare presented high sensitivity & specificity for diagnosing thyroid diseases
- Strong correlations were observed between Finecare and the reference method
- Sample types (serum, venepuncture, fingerstick) do not affect TSH Finecare results
- Fingerstick/capillary sample is appropriate for TSH quantitation by Finecare

## 1. Introduction

Endocrine and metabolic disorders are common worldwide [1-3]. Among these, thyroid dysfunction remains a major problem [1]. The measurement of thyroid-stimulating hormone (TSH) levels represents the first-line assay for assessing thyroid function [2]. TSH is the most significant test for understanding any relevant thyroid problems [3]; along with triiodothyronine (T3) and thyroxine (T4) testing, primary or secondary thyroid disease can be diagnosed [4].

TSH reference range values are affected by age and gender [5]. However, according to the literature, the normal/reference range for male and female adults is generally between 0.35 to 4.5 mIU/L [9-10][3]. Values above 4.5 mIU/L indicate low thyroid function (hypothyroidism), and values less than 0.35 mIU/L indicate hyperactive thyroid [8]. The frequency of thyroid dysfunction cases dictates the need for skilled physicians to diagnose thyroid diseases [9]. Another prerequisite for thyroid dysfunction diagnoses and follow-up treatments is to provide reliable analytical equipment for testing thyroid-related hormones. Because TSH is the first-line test to screen or confirm thyroid disease, equipment with high sensitivity and specificity is always needed to reduce the incidence of false diagnoses. Failure to assess thyroid dysfunction puts the patient at high risk for different conditions, such as infertility, osteoporosis, and cardiovascular disease [10].

Immunoradiometric assay (IRMA) is considered the most sensitive assay and a reference method for quantitative measurement of many analytes and hormones, including TSH [11]. Due to many limitations of the IRMA, the third generation fully automated chemiluminescence or electrochemiluminescence immunoassays (CLIA or ECLIA) became the most popular and sensitive assays to measure hormones [12-14]. For instance, the Elecsys’ fully automated ECLIA system, such as Cobas, which developed by Roche, is now considered one of the most reliable systems for analyzing TSH with clinical suspicion of thyroid disease [12-14].

The demand for rapid measuring of results (e.g., in intensive care and newborn screening) and the development of a testing method that can also be operated, by nursing staff, with minimal cost are factors contributing to the increasing use of point-of-care testing (POCT) equipment. While comparable with conventional laboratory assays (ELISA and CLIA) in terms of sensitivity and specificity, the POC tests, such as lateral flow immunoassays (LFIAs) save time and reduce costs [15]. Recently many commercial LFIAs for measuring TSH were developed by different manufacturers. However, most of these LFIAs are mainly used for screening primary hypothyroidism in children (only detects values more than 5 mIU/L) and are not approved in the USA or Europe [16]. The issues with those methods are the lack of sensitivity and accuracy and that they cannot measure a wide range of readings. Additionally, most LFIA use plasma or serum, which requires extensive processing and trained personnel to handle the blood withdrawal and sample preparation for analysis [16].

In the last 15 years, many LFIAs were developed and proposed for quantitative measurement of TSH[17][18].However, their performance remains to be compared to a standard and reliable laboratory assay. Therefore, in the present study, we aim to evaluate the performance of the Finecare™ TSH Rapid Quantitative Test using samples obtained from fingerstick/capillary, venepuncture whole-blood and serum. We used the Elecyes system Roche CobasPro-c503 ECLIA analyzer from Roche Diagnostics as a reference method for this evaluation. In addition, we aim to evaluate the effect of using different type of samples (serum, fingerstick and whole-blood) on Finecare™ TSH quantitative results.

## 2. Materials and Methods

### 2.1 Sample collection and ethical approval

In this study, a 102 fingersticks/capillary and matched venepuncture whole-blood, and serum samples were collected from participants visiting a clinical laboratory in Jordan. Data were collected in 2022. Ethical approval was granted for data collection by Qatar University (IRB#. QU-IRB 1766-E/22).

### 2.2 Finecare™ TSH rapid quantitative test

The Finecare™ TSH relies on a solid sandwich immunodetection method to rapidly quantify TSH. Fingerstick or EDTA whole-blood and coagulated fresh blood samples were processed following the manufacturer’s recommendations. Briefly, 75 μL of fingerstick, venepuncture whole-blood or serum was added to the detection buffer tube. The sample and the buffer were mixed well, then 75 μL of the mixture was loaded into the sample well of the test device. The sample was inserted into the sample holder of the Finecare™ FIA meter. The reaction time was 15 minutes. The deviations beyond the reference range of 0.35 to 4.5 mIU/L were considered positive [6].

### 2.3 Reference method Roche CobasPro-c503

The Elecsys® Anti-TSHR test uses anti-human THS monoclonal antibodies labelled with ruthenium complex. The test follows a sandwich principle and takes around 18 minutes for the complete execution. All samples were processed following the manufacturer’s recommendations. Briefly, 50 μL of the patient’s sample was incubated with the TSH-specific monoclonal antibody labelled with a ruthenium complex, which will react to form a sandwich complex. After that, streptavidin-coated microparticles were added, and samples were incubated. During the incubation, the complex binds to the solid phase via biotin and streptavidin interaction. The reaction mixture is added to the measuring cell, so the microparticles are captured into the electrode surface by magnetic attraction. Following the additions of ProCell M solution to remove the unbound substances, a voltage is applied to the electrode, which induces chemiluminescent emission measured by a photomultiplier.

### 2.4 Statistical analysis

Data were analyzed using GraphPad Prism 9.3.1 (San Diego, CA, USA). The collected dataset was subjected to the Shapiro-Wilk normality test to evaluate data normality. In the absence of normal distribution, non-parametric tests were performed using Kruskal-Wallis to test for the differences between independent samples. p-values ≤0.05 were considered statistically significant.

Using Roche CobasPro-c503 as the reference standard, concordance analysis based on 2×2 contingency tables was conducted. These concordance measures included overall percentage agreement (OPA), positive (PPV), and negative predictive values (NPV), as well as Cohen’s Kappa statistics. The latter measure is a standard and robust metric that estimates the level of agreement, beyond chance, between two diagnostic tests. Ranging between 0 and 1, a Cohen’s Kappa value <0.40 denotes poor agreement, 0.40-0.59 denotes fair agreement, 0.60-0.74 denotes good agreement, and ≥0.75 denotes excellent agreement [19]. The significance level was indicated at 5%, and a 95% confidence interval (CI) was reported for each metric. Correlation and linear regression analysis were performed between Finecare™ and the reference method, and between Finecare™’s different blood draws. Spearman correlation coefficient (r) was calculated. For absolute values of spearman’s r, 0-0.19 is denoted as a very weak correlation, 0.2-0.39 as weak, 0.40-0.59 as moderate, 0.6-0.79 as strong, and 0.8-1 indicates a very strong correlation. Confidence interval (CI) at 95% were indicated for all tests [20].

## 3. Results

### 3.1 Sample type (fingerstick, venepuncture whole-blood, or serum) does not significantly affect the results obtained by Finecare™

Finecare™ performance was assessed by comparing the Finecare™ fingerstick, venepuncture, and serum samples to the same serum samples analyzed by Roche CobasPro-c503. The general distribution for all numerical values obtained by Finecare™ and the reference method is represented in Figure 1. There is no significant difference between the overall values and medians obtained by Fincare™ as compared with Cobas Pro-c503 (Figure 1). These results suggest that sample type has no significant effect on the obtained results (Figure 1).

**Figure 1.**
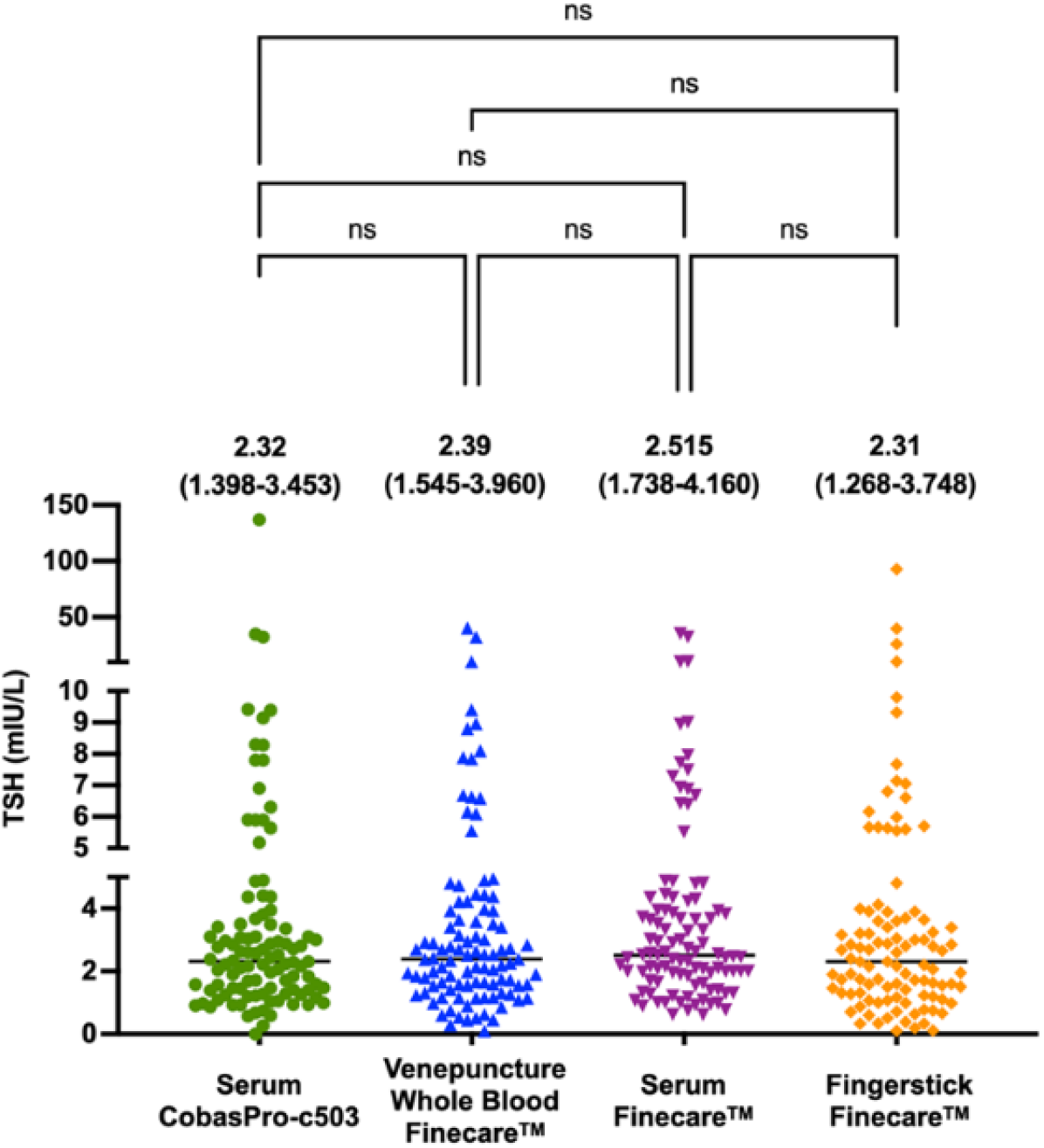
General distribution of values obtained from fingerstick, venepuncture whole-blood and serum using Finecare™ machine and the reference method, Roche CobasPro-c503. The difference between all groups was obtained using the nonparametric Kruskal-Wallis test. The median and the interquartile ranges are presented above each test. ns, non-significant (p>0.05)

### 3.2 Finecare™ quantitative results are highly correlated with the reference method

We performed a correlation analysis between Roche CobasPro-c503 reference method and Finecare™ TSH test values of 102 samples. As indicated in figure 2, both Finecare™ venepuncture whole-blood and fingerstick sample test values have a very strong correlation with Roche CobasPro-c503 (r=0.95 and r=0.9 respectively; p<0.0001). Strong correlation was also obtained between Finecare™ serum sample test values and CobasPro-c503 (r=0.93, p<0.0001) (Figure 3).

**Figure 2.**
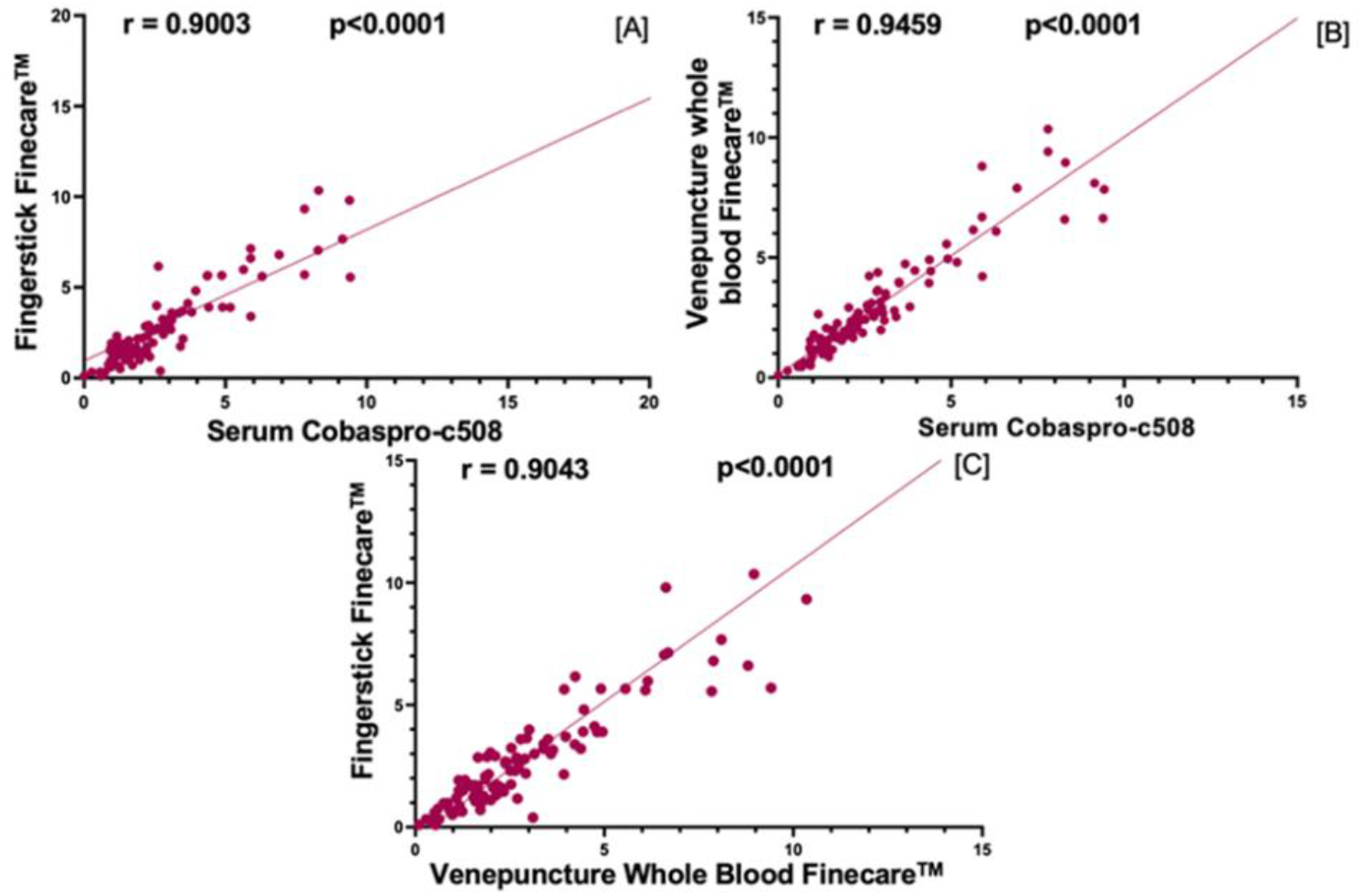
Pairwise correlation analysis and linear regression analysis of the numerical values obtained by each assay. Spearman correlation coefficient (r) was calculated to be 0.9003, 0.9459, and 0.9043 for [A], [B], and [C], respectively. P values are indicated (<0.0001).

**Figure 3.**
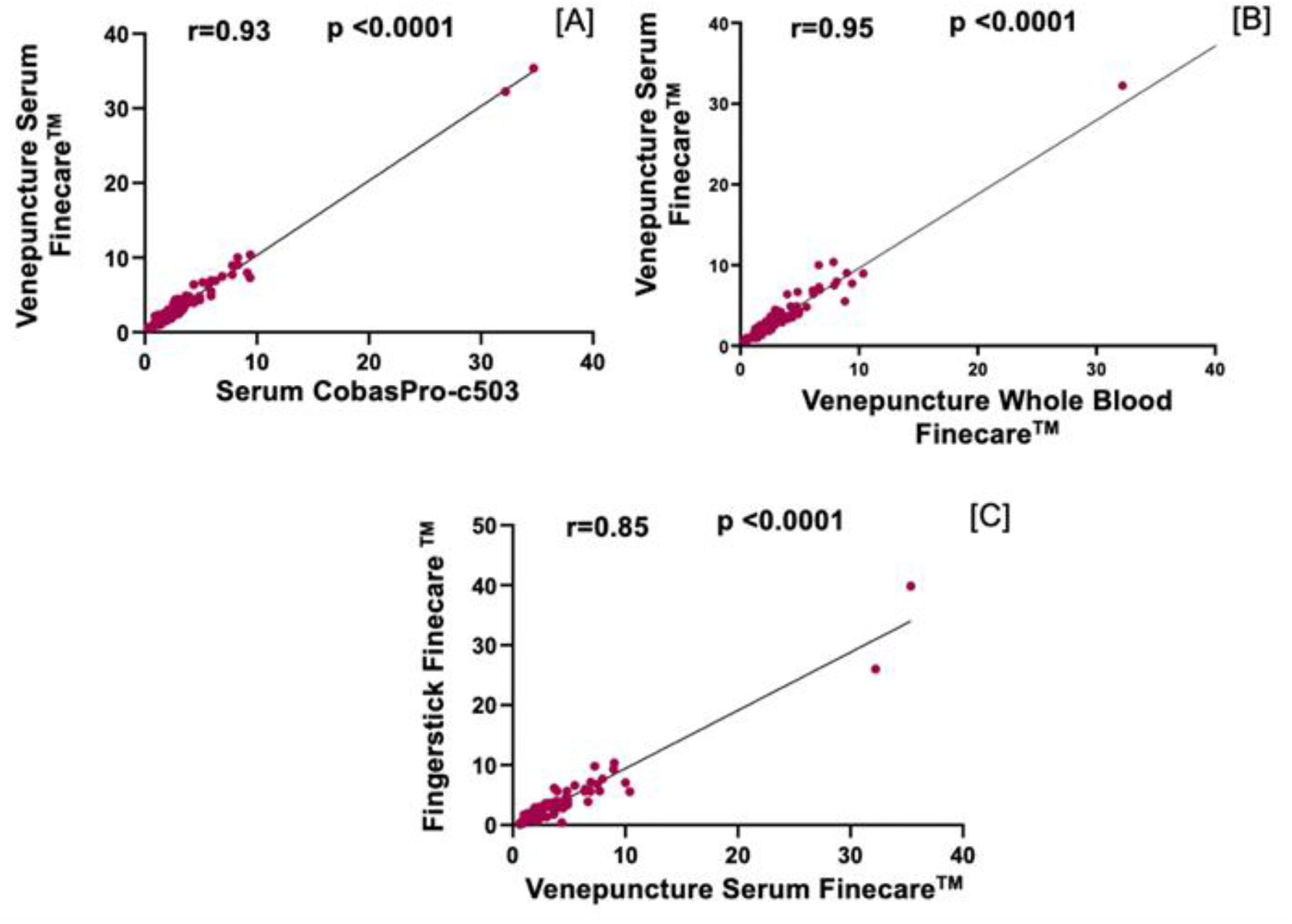
Pairwise correlation and linear regression analyses of the numerical values obtained by each assay. Spearman correlation coefficient (r) was calculated to be 0.93, 0.95, and 0.85 for [A], [B], and [C], respectively. P values are indicated (<0.0001).

Finecare™ venepuncture whole-blood and serum samples showed a strong correlation (r=0.95, p<0.0001). Finecare™ fingerstick showed similar results when compared to the serum sample as in the venepuncture whole-blood samples (r=0.85, p<0.0001) (Figure 3). These results confirm that the sampling method (i.e., serum, vs fingerstick vs venepuncture) does not affect the TSH test values, and suggest that Fincare™ can be used for analytical quantitation of TSH regardless of the sampling method.

### 3.3. Finecare™ showed high true positive and negative rates compared to the reference method

We classified the disease condition according to the test value obtained by the reference method (Table 1). Results are considered positive (diseased) if any of TSH values were below 0.35 or above 4.5 mIU/mL. TSH serum, venepuncture, and fingerstick results were also classified accordingly. As shown in Table 2, the positive and negative values of Finecare™ were comparable to those of the reference method, regardless of the types of samples (with very few exceptions). We believe these exceptions are due to the differences between Finecare and reference method borderline results, which are outside, yet very close, to the upper or lower limit of the normal/reference range (0.35-4.54 mIU/mL).

**Table 1.**
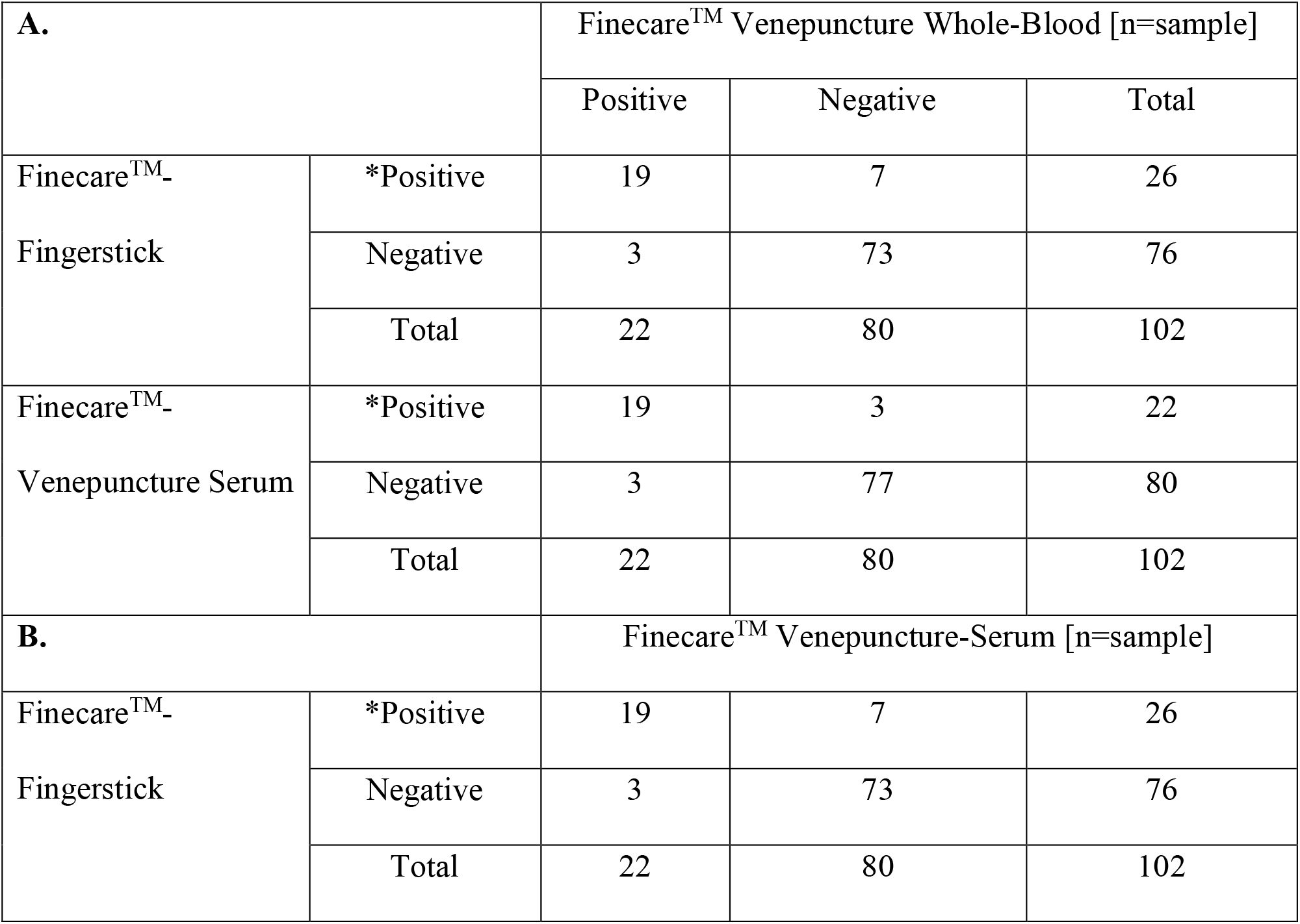
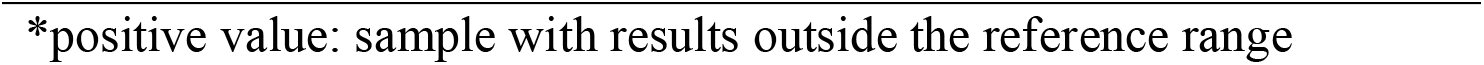
A comparison between the three Finecare™ testing methods: venepuncture whole-blood, venepuncture serum, and fingerstick.

| A. |  | Finecare™ Venepuncture Whole-Blood [n=sample] |  |  |
| --- | --- | --- | --- | --- |
|  |  | Positive | Negative | Total |
| Finecare™<br>Fingerstick | *Positive | 19 | 7 | 26 |
|  | Negative | 3 | 73 | 76 |
|  | Total | 22 | 80 | 102 |
| Finecare™<br>Venepuncture Serum | *Positive | 19 | 3 | 22 |
|  | Negative | 3 | 77 | 80 |
|  | Total | 22 | 80 | 102 |
| B. |  | Finecare™ Venepuncture-Serum [n=sample] |  |  |
| Finecare™<br>Fingerstick | *Positive | 19 | 7 | 26 |
|  | Negative | 3 | 73 | 76 |
|  | Total | 22 | 80 | 102 |

**Table 2.**
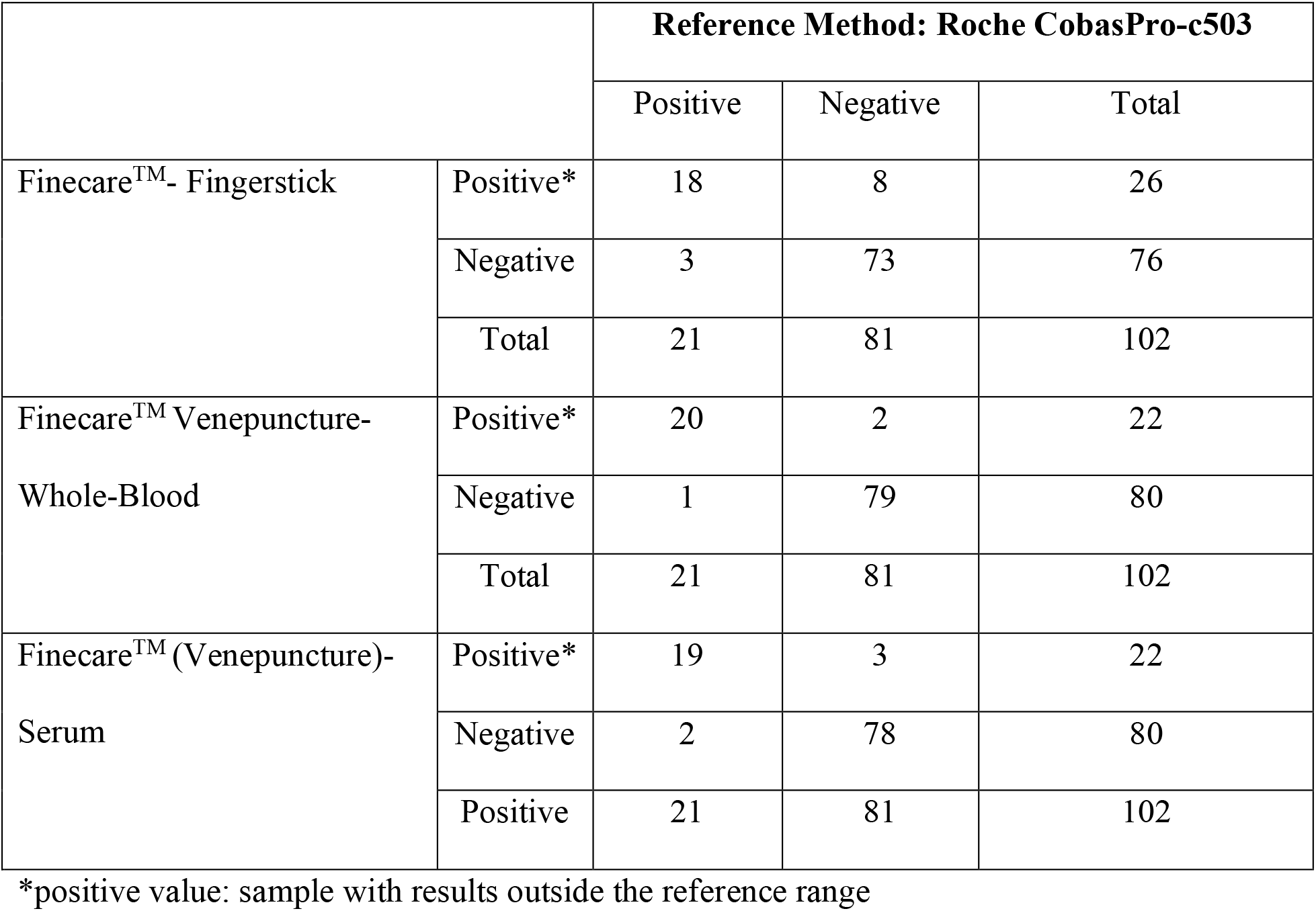
A comparison between the three Finecare™ testing methods (venepuncture whole-blood, venepuncture serum, and fingerstick) against the reference

|  |  | Reference Method: Roche CobasPro-c503 |  |  |
| --- | --- | --- | --- | --- |
|  |  | Positive | Negative | Total |
| Finecare™- Fingerstick | Positive* | 18 | 8 | 26 |
|  | Negative | 3 | 73 | 76 |
|  | Total | 21 | 81 | 102 |
| Finecare™ Venepuncture- Whole-Blood | Positive* | 20 | 2 | 22 |
|  | Negative | 1 | 79 | 80 |
|  | Total | 21 | 81 | 102 |
| Finecare™ (Venepuncture)- Serum | Positive* | 19 | 3 | 22 |
|  | Negative | 2 | 78 | 80 |
|  | Positive | 21 | 81 | 102 |

\*positive value: sample with results outside the reference range

### 3.4 Finecare™ has high sensitivity and specificity for diagnosis of thyroid abnormalities

To further confirm our results, a concordance analysis between Finecare™ venepuncture whole-blood and Finecare™ serum was performed (Table 3). The serum Finecare™ was considered the reference for comparison. The Finecare™ venepuncture whole-blood showed high overall percentage agreement (OPA) of 94.1% (86.4%-96.3%), positive predictive value (PPV) of 86.4% (67.3%-95.1%), negative predictive value (NPV) of 96.25% (90%-98.7%) and an excellent agreement with the Finecare™ serum [Cohen’s Kappa=0.826 (0.692-0.960)]. If Finecare™ serum considered as a reference method, the sensitivity and specificity of Finecare™ venepuncture were 86.4% (65.1%-97.1%) and 96.3% (89.4%-99.2%), respectively (Table 3). Moreover, Finecare™ venepuncture whole-blood showed high sensitivity and specificity compared to the reference method, 95.2% and 97.5%, respectively (Table 3). Finecare™ serum samples showed a high sensitivity of 95.1% and a specificity of 90.5%, yet slightly lower than the venepuncture whole-blood samples when both, serum and venepuncture, were compared to the reference method.

**Table 3.**
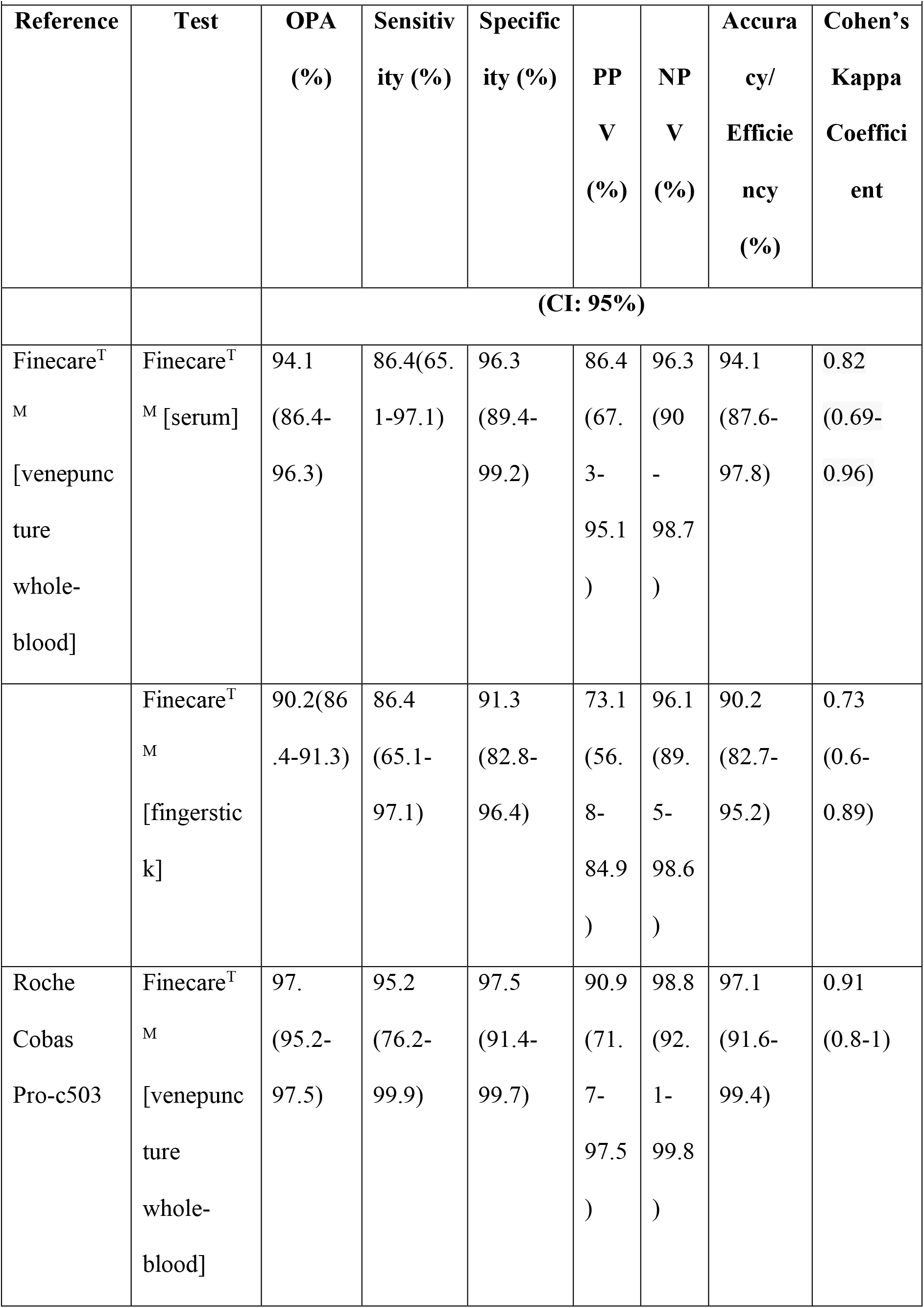
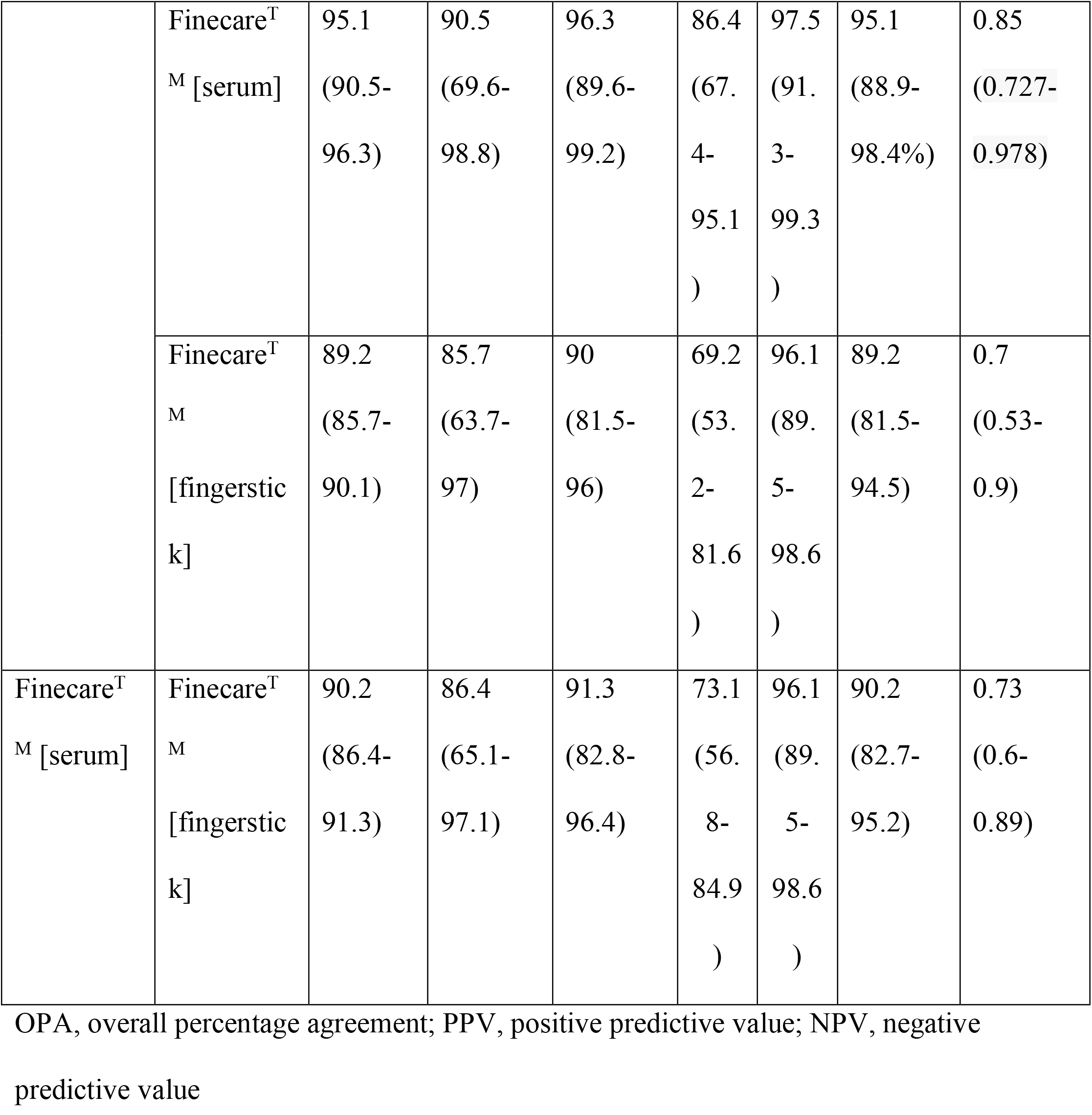
Validity, agreement, and accuracy of Finecare in comparison with the reference method

Finecare™ fingerstick also showed good performance but with a lower sensitivity (85.7%) and specificity (90%) than the serum and the venepuncture whole blood results (Table 3). When comparing Finecare™ venepuncture whole-blood and serum samples to fingerstick, similar results were obtained with a sensitivity of 90.2% and specificity of 86.4% (Table 3). Moreover, different test agreements and Cohen’s Kappa statistics were reported between the reference and Finecare™ venepuncture and fingerstick samples. The OPA, PPV, and NPV between the reference method and venepuncture blood were high: 97.1%, 90.9% and 98.8%, respectively. Comparing Finecare™ fingerstick to the reference method, OPA, PPV, and NPV were lower than those of the venepuncture blood samples; 89.2%, 69.2% and 96.1%, respectively. A good agreement was found between the Finecare™ fingerstick and the reference method (Cohen’s Kappa=0.7) and an almost perfect agreement between the Finecare™ venepuncture and the reference (Cohen’s Kappa=0.91). These results indicate that Fincare™ TSH can be used as a qualitative screening of TSH abnormalities regardless the source of the specimen withdrawal.

## Discussion

Congenital hypothyroidism results from the failure of the thyroid gland to produce adequate levels of the thyroid hormones [21], a condition that requires an immediate diagnosis, especially in newborns. In this regard, the rapid turnaround time, minimal sample volume, and elimination of hematocrit bias make POCT TSH assays the method of choice for newborn thyroid screening [16]. Moreover, POCT TSH assay affordability and ease of use enable thyroid diagnostic testing in resource-limited settings [22].

So far, many POCTs have been developed; however, their performance remains to be compared to standard laboratory approaches. In this study, Finecare™ TSH Rapid Quantitative Test performance was validated along with Finecare™ FIA System for the quantitative determination of TSH in human blood. This test is used as POCT for screening and following up on TSH values in the population. This study employed a total of 102 samples to evaluate the assays’ performance. To our knowledge, this is one of the first studies conducted to validate the fluorescence-LFIA-based Finecare™ TSH test, besides the one by Kahaly et al. 2022 where the performance of the Finecare™ TSH test was compared to Abbott [23].

The presented data demonstrate that Finecare™ results are consistent with the reference laboratory method (Roche CobasPro-c503) using venepuncture whole-blood, serum and fingerstick samples. Venepuncture whole-blood samples showed excellent sensitivity and specificity, 95.2% and 97.5% (Table 3). Excellent agreement between the two tests was also observed (Cohen’s Kappa=0.91) along with a very strong correlation (r=0.95, p<0.0001) (Table 3, Figure 2). In contrast, Finecare™ serum samples showed slightly less sensitivity and specificity (90.5% and 96.3%, respectively) (r=0.93, p<0.0001). This is mainly due to borderline results but not due to significant differences between the values. Since the reference range is set between 0.35 to 4.5 mUI/L, any value outside this range, even with one decimal point, is considered positive, therefore affecting the overall test sensitivity. Although in this study we considered 0.35 and 4.5 mUI/L as the reference range, others laboratories use different ranges (based on the population origin) [22-23], which is an aspect that results in different sensitivity and specificity values. Nonetheless, correlation results indicate a very strong agreement suggesting that Finecare™ is suitable for quantitative TSH measurement. Furthermore, the Finecare™ fingerstick showed 85.7% sensitivity and 90% specificity, fair test agreement (Cohen’s Kappa=0.7) and very strong correlation (r=0.9, p<0.0001). The excellent concordance between the POCT Finecare™ and Roche CobasPro-c503 makes it an attractive alternative to the standard laboratory technique in a non-laboratory setting. This has been challenging because almost all of the currently available TSH POCTs were reported to have either high sensitivity but low specificity or vice versa. For instance, TSH-CHECK-1 © (Vedalab, Alençon, France) test sensitivity was 100.0%, but specificity was 76.6% [26], indicating high false-positive results, which can possibly be due to cross-reactivity. It is noteworthy to mention that, due to hematocrit variability, it is generally believed that serum or plasma TSH assays are more accurate, affordable, accessible, and clinically useful than tests which measure TSH in a capillary or venepuncture whole-blood sample [27].

There are great advantages to using Finecare™ as POCT since results can be obtained in a short period of time, and there is no need for lengthy sample processing since capillary blood can be used (easy to collect with small volumes), especially for neonates. Fingerstick samples analyzed by Finecare™ showed a very strong correlation with the reference method (r=0.9), making it feasible for TSH screening and following up on quantitative measurement.

There are a few variations between the data obtained with Finecare™ fingerstick and the reference method. Such variations are primarily due to technical errors rather than instrumental faults, which could be attributed to incorrect fingerstick sampling or sample clotting at the time of sample collection. As a result, sample clotting could occur. The collection tube of Wondfo TSH kit does not have heparin; thus, partial clotting in the sample could affect the results. Additionally, other issues could derive from mistakes or inaccuracies in volume pipetting when withdrawing the fingerstick samples since the test requires the handling of relatively large volumes from the tip of the finger (75µL). Variations in fingerstick and CobasProc-c503 results could also happen due to artifact from skin contaminants during sample withdrawal. Finally, variations between assays are expected since the used test principles are different. In this regard, studies had reported variabilities in values also when similar test principles such as CLIA and ELISA were used [28]. Discrepancies in values obtained by Roche Cobas and Abbott TSH test were reported [29], although both companies are considered leading CLIA manufacturers worldwide. In conclusion, Finecare™ is a reliable assay and can be easily implemented for screening and following up TSH values in the population, particularly in none or small laboratory settings. Finecare™ could yield more accurate results if the company would reduce the sample volume and would implement the use of heparinized tubes for blood withdrawal.

## Data Availability

All data produced in the present study are available upon reasonable request to the authors

## Conflict of Interest

GKN is currently an associate editor at Heliyon Infectious Disease Journal.

MMA would like to acknowledge that he received the Finecare TSH kit and reader as an in-kind support for this study.

## Acknowledgement

MMA would like to acknowledge his lab staff for the technical support.

## Funding Source

None

## Notes

### Funding Statement

This study did not receive any funding

### Author Declarations

Ethics committee/IRB of Qatar University waived ethical approval for this work.

## References

[1] P. N. Taylor et al., “Global epidemiology of hyperthyroidism and hypothyroidism,” Nat. Rev. Endocrinol., vol. 14, no. 5, pp. 301–316, 2018, doi: 10.1038/nrendo.2018.18.

[2] A. Di Cerbo, N. Quagliano, A. Napolitano, F. Pezzuto, T. Iannitti, and A. Di Cerbo, “Comparison between an emerging point-of-care tool for TSH evaluation and a centralized laboratory-based method in a cohort of patients from southern Italy,” Diagnostics, vol. 11, no. 9, 2021, doi: 10.3390/diagnostics11091590.

[3] M. T. Sheehan, “Biochemical testing of the thyroid: TSH is the best and, oftentimes, only test needed - A review for primary care,” Clin. Med. Res., vol. 14, no. 2, pp. 83– 92, 2016, doi: 10.3121/cmr.2016.1309.

[4] D. B. Dunlap, “Dickson_1990_Book on Thyroid function,” in Clinical Methods: The History, Physical, and Laboratory Examinations, 3rd editio., E. In: Walker HK, Hall WD, Hurst JW, Ed. Boston: Boston: Butterworths, 1990, pp. 666–676. [Online]. Available: https://www.ncbi.nlm.nih.gov/books/NBK249/

[5] X. Chen et al., “Relationship of gender and age on thyroid hormone parameters in a large chinese population,” Arch. Endocrinol. Metab., vol. 64, no. 1, pp. 52–58, 2020, doi: 10.20945/2359-3997000000179.

[6] A. Allahabadia, “Autoimmune Hypothyroidism with Persistent Elevation of TSH,” in A Case-Based Guide to Clinical Endocrinology, T. F. (eds) A. C.-B. G. to C. E. Davies, Ed. Humana Press, 2008, pp. 85–91. doi: 10.1007/978-1-60327-103-5_9.

[7] G. Targher et al., “Association between serum TSH, free T4 and serum liver enzyme activities in a large cohort of unselected outpatients,” Clin. Endocrinol. (Oxf)., vol. 68, no. 3, pp. 481–484, 2008, doi: 10.1111/j.1365-2265.2007.03068.x.

[8] J. R. Garber et al., “Clinical practice guidelines for hypothyroidism in adults: Cosponsored by the american association of clinical endocrinologists and the American thyroid association,” Endocr. Pract., vol. 18, no. 6, pp. 988–1028, 2012, doi: 10.4158/ep12280.gl.

[9] A. Goel, C. Shivaprasad, A. Kolly, A. A. Pulikkal, R. Boppana, and C. S. Dwarakanath, “Frequent occurrence of faulty practices, misconceptions and lack of knowledge among hypothyroid patients,” J. Clin. Diagnostic Res., vol. 11, no. 7, pp. OC15–OC20, 2017, doi: 10.7860/JCDR/2017/29470.10196.

[10] D. Mendes, C. Alves, N. Silverio, and F. B. Marques, “Prevalence of Undiagnosed Hypothyroidism in Europe: A Systematic Review and Meta-Analysis,” Eur. Thyroid J., vol. 8, no. 3, pp. 130–143, 2019, doi: 10.1159/000499751.

[11] R. G. Bretzel, G. Siedenberg, E. Skribelka, H. Schatz, and K. Federlin, “Application of a sensitive immunoradiometric assay (IRMA) for T S H in the diagnosis and follow-up thyroid disorders,” 1987.

[12] M. Sánchez-Carbayo, M. Mauri, R. Alfayate, C. Miralles, and F. Soria, “Analytical and clinical evaluation of TSH and thyroid hormones by electrochemiluminescent immunoassays,” Clin. Biochem., vol. 32, no. 6, pp. 395–403, 1999, doi: 10.1016/S0009-9120(99)00032-6.

[13] F. Kazerouni and H. Amirrasouli, “Performance characteristics of three automated immunoassays for thyroid hormones,” Casp. J. Intern. Med., vol. 3, no. 2, pp. 400–404, 2012.

[14] R. Sarkar, “TSH comparison between chemiluminescence (architect) and electrochemiluminescence (Cobas) immunoassays: An Indian population perspective,” Indian J. Clin. Biochem., vol. 29, no. 2, pp. 189–195, 2014, doi: 10.1007/s12291-013-0339-7.

[15] K. M. Koczula and A. Gallotta, “Lateral flow assays,” Essays Biochem., vol. 60, no. 1, pp. 111–120, 2016, doi: 10.1042/EBC20150012.

[16] T. Association, “Point-of-Care Thyroid Diagnostics and Thyroid Disease Management Point-of-Care Thyroid Diagnostics and Thyroid Disease Management,” pp. 2018–2020, 2019.

[17] Y. Yazawa, T. Oonishi, K. Watanabe, A. Shiratori, S. Funaoka, and M. Fukushima, “System-on-fluidics immunoassay device integrating wireless radio-frequency-identification sensor chips,” J. Biosci. Bioeng., vol. 118, no. 3, pp. 344–349, 2014, doi: 10.1016/j.jbiosc.2014.02.010.

[18] J. Ylikotila, L. Välimaa, M. Vehniäinen, H. Takalo, T. Lövgren, and K. Pettersson, “A sensitive TSH assay in spot-coated microwells utilizing recombinant antibody fragments,” J. Immunol. Methods, vol. 306, no. 1–2, pp. 104–114, 2005, doi: 10.1016/j.jim.2005.08.002.

[19] E. In: Kirch W, Ed., “Kappa Coefficient Kappa coefficient,” Dordrecht: Springer Netherlands: Encyclopedia of Public Health., 2008.

[20] D. Middleton, “‘ Statistics at square one,’ “ Br. Med. J., vol. 2, no. 6042, pp. 1008–1009, 1976, doi: 10.1136/bmj.2.6042.1008-b.

[21] M. V Rastogi and S. H. Lafranchi, “Congenital hypothyroidism,” pp. 1–22, 2010.

[22] J. Fualal and J. Ehrenkranz, “Access, availability, and infrastructure deficiency : The current management of thyroid disease in the developing world,” Springer Sci., pp. 583–589, 2016, doi: 10.1007/s11154-016-9376-x.

[23] G. J. Kahaly et al., “A novel point-of-care device accurately measures thyrotropin in whole blood, capillary blood and serum,” Clin. Chem. Lab. Med., vol. 60, no. 10, pp. 1607–1616, 2022, doi: 10.1515/cclm-2022-0525.

[24] R. Meamar et al., “Thyroid stimulating hormone reference range: Iranian thyroid cohort study,” Acta Biomed., vol. 92, no. 5, pp. 1–10, 2021, doi: 10.23750/abm.v92i5.9643.

[25] R. Turkal et al., “Accurate interpretation of thyroid dysfunction during pregnancy: should we continue to use published guidelines instead of population-based gestation-specific reference intervals for the thyroid-stimulating hormone (TSH)?,” BMC Pregnancy Childbirth, vol. 22, no. 1, pp. 1–14, 2022, doi: 10.1186/s12884-022-04608-z.

[26] C. S. Kosack, A. Page, L. T. Van Hulsteijn, and E. G. W. M. Lentjes, “TSH-CHECK-1 Test : Diagnostic Accuracy and Potential Application to Initiating Treatment for Hypothyroidism in Patients on Anti-Tuberculosis Drugs,” vol. 7, no. 3, pp. 7–10, 2012, doi: 10.1371/journal.pone.0033704.

[27] E. M. Hall, S. R. Flores, N. Screening, and M. B. Branch, “HHS Public Access,” Int J Neonatal Screen., vol. 1, no. 2, pp. 69–78, 2017, doi: 10.3390/ijns1020069.Influence.

[28] A. History and M. E. M. Gar-elnabi, “Assessment of human thyroid function using radioimmunoassay and enzyme-linked-,” 2013, doi: 10.5835/jecm.omu.30.04.007.

[29] T. Kalaria et al., “The diagnosis and management of subclinical hypothyroidism is dependent – Implications for clinical practice,” no. December 2020, pp. 1012–1016, 2021, doi: 10.1111/cen.14423.

